# Effects of statins on lipid profile of kidney transplant recipients: a meta-analysis of randomized controlled trials

**DOI:** 10.1101/2020.04.07.20057349

**Authors:** Xiu Huang, Yong Jia, Xiaoyu Zhu, Yangyang Zhang, Lili Jiang, Xuejiao Wei, Dan Zhao, Xiaoxia Zhao, Yujun Du

## Abstract

**Objective:** To assess the benefits of statins on lipid profile in kidney transplant recipients via a meta-analysis.

**Methods:** We systematically identified peer-reviewed clinical trials, review articles, and treatment guidelines from PubMed, Embase, the Cochrane Library, Wanfang, Chinese National Knowledge Infrastructure (CNKI), SinoMed (CBM), and Chongqing VIP databases from inception to April 2019. In the analysis, only randomized controlled clinical trials performed in human were included.

**Results:** Eight articles were included in the analysis, involving 335 kidney transplant recipients who received statins and 350 kidney transplant patients as the control group. Results revealed that statins improved the lipid profile of kidney transplant recipients. Specifically, statin therapy significantly reduced total cholesterol and low-density lipoprotein cholesterol. However, it had no effects on high-density lipoprotein cholesterol and triglycerides levels.

**Conclusions:** The present study provides valuable knowledge on the potential benefits of statins in kidney transplant recipients. This meta-analysis shows that statin therapy modifies the lipid profile in this patient population.

## 1. Introduction

The burden of chronic kidney disease (CKD) is rapidly increasing worldwide. Studies have reported that cardiovascular disease (CVD) is a leading cause of death in patients with CKD, especially kidney transplant recipients(KTRs)1. It has been estimated that the mortality rate of CVD is 17-50% in KTRs2. Previously, it was found that there are many risk factors contributing to CVD. Among the important risk factors for cardiovascular disease are high level of atherosclerosis, ageing, heightened systemic inflammation, oxidative stress, endothelial dysfunction, hypertension, insulin resistance and diabetes mellitus, cigarette smoking and dyslipidemia. Data show that dyslipidemia is one of the most common metabolic complications and the incidence of it is as high as 80% in KTRs3.

Dyslipidemia is not only associated with high CVD risk, but also a independent risk factor for allograft rejection and graft survival in KTRs4. Aware of the harm of post-transplant dyslipidemia and optimize suitable therapeutic strategies are crucial to KTRs. Therefore, it’s reasonable to assume that the intervention of dyslipidemia could improve lipid profile in KTRs.

Statins (inhibitors of 3-hydroxy-3-methylglutaryl coenzyme A (HMG-CoA) reductase) may be a good choice for the primary and secondary prevention drugs of CVD and the treatment of lipid profile. There are a series of studies that have focused on lipid profile in all CKD, however, few studies are about KTRs that an overlooked group and there are well discrepancies between the results of these studies. For example, Palmer 5showed that statins can significantly reduce serum total cholesterol(TC),low-density lipoprotein cholesterol (LDL-C), lowered high-density lipoprotein cholesterol(HDL-C) and serum triglycerides (TG). However, Messow 6 held that lowering lipid about statins was not effective in patients after renal transplant.

The reasons that we do not replicate these studies are as follows: firstly, it remains controversial about the benefits of statins on lipid profile in kidney transplant recipients so far; and secondly, in the last few years, there have been published some original studies about it. Lastly, we have already searched the Chinese database that were different from other meta-analysis. Thus, this meta-analysis is aiming to critically appraise scientific evidence about the efficacy of statins on lipid profile in KTRs.

## 2. Methods

### 2.1. Eligibility criteria

Only RCTs were inclusive within this review. Cross-sectional studies, cohorts, case studies and cross-over experiments were excluded to be able to determine the effectiveness of statins intervention. The study population was adults (more than 18 years) who have been only received kidney transplant without multiorgan transplant for over 1 month. The intervention was statins therapy involving all kinds of statins. The control group comprised patients receiving placebo, routine care and no treatment. The primary outcome was serum lipid level including TC, LDL-C, HDL-C and TG.

### 2.2 Data sources and search strategy

All publications were searched on English databases which included the PubMed, Cochrane Library, Embase and the Chinese databases which included the Wanfang database, Chinese National Knowledge Infrastructure (CNKI), VIP database, and SinoMed from inception to April 2019 without language restriction.

The following search terms were used: renal transplantation, kidney transplantation, acute renal allograft rejection, renal allograft rejection, hypercholesterolaemia, hyperlipidaemia, dyslipidaemia, hydroxymethylglutaryl coenzyme a reductase inhibitor,HMG-CoA reductase inhibitors, hydroxymethylglutaryl-CoA reductase inhibitors, statin, pravastatin, fluvastatin, lovastatin, simvastatin, atorvastatin, cerivastatin and rosuvastatin. Further details of the search strategies are available from the authors by request. The reference lists of identified trials and review articles were scrutinized for additional trials. Draft Embase and SinoMed search strategies are included as Supplementary Appendix 1. All studies included in the meta-analysis were randomized controlled clinical trials.

### 2.3. Study selection

Eligible studies were identified by screening the titles and abstracts. Two reviewers (Miss Huang and Miss Zhu) independently screened all databases to retrieve studies that met the eligibility criteria. Disagreements on whether an article was eligible were resolved by discussion or were arbitrated by a third reviewer (Mr. Jia).

### 2.4. Data extraction

The following data were extracted from eligible trials: author’s name, year of publication, types of statins, duration, the dose of statins, the number of total patients, the duration of follow-up, the level of TC, LDL-C, HDL-C and TG after treatment. The extraction results were re-analyzed by two independent reviewers. Discrepancies were solved by discussion or through re-evaluation by a third reviewer.

### 2.5. Risk of bias

The Cochrane Collaboration’s tool was utilized to assess the bias risk in all the included studies. The following sources of bias were recorded: method of randomization, allocation concealment, blinding of participants and personnel, binding of outcome assessment, incomplete outcome data, selective reporting bias and other bias. Each article was categorized either as ‘high risk’, ‘low risk’, or ‘unclear risk’, with the last category meaning that either lack of information or unclear about the potential for bias.

### 2.6. Statistical analysis

Included studies were weighted by effect size and pooled. The Review Manager software 5.3 was used to assess standardized mean differences (SMDs) with 95% confidence intervals (95%CIs) which were calculated using Der Simonian and Laird (for random effects) methods. The Cochran Q test and the I^2^ statistic with 95% CIs were performed to determine the level of heterogeneity7 and statistical significance was defined as p<0.05. Given that an I^2^ equaling 0 does not exist, values of 95% CIs ≥ 50% were used to represent high heterogeneity 8. The random-effects model was applied in the meta-analysis. Funnel plots that were calculated in the Review Manager software were utilized to indicate publication bias. Subgroup analyses were not performed because of the limited number of studies.

## 3. Results

### 3.1. Study selection

The search identified 7278 records of which 2030 potentially relevant full articles were found to be eligible (Figure 1). Finally, eight studies met the inclusion criteria 9-16, covering 350 patients who received kidney transplant, and 335 patients randomized to the control group.

**Fig. 1.**
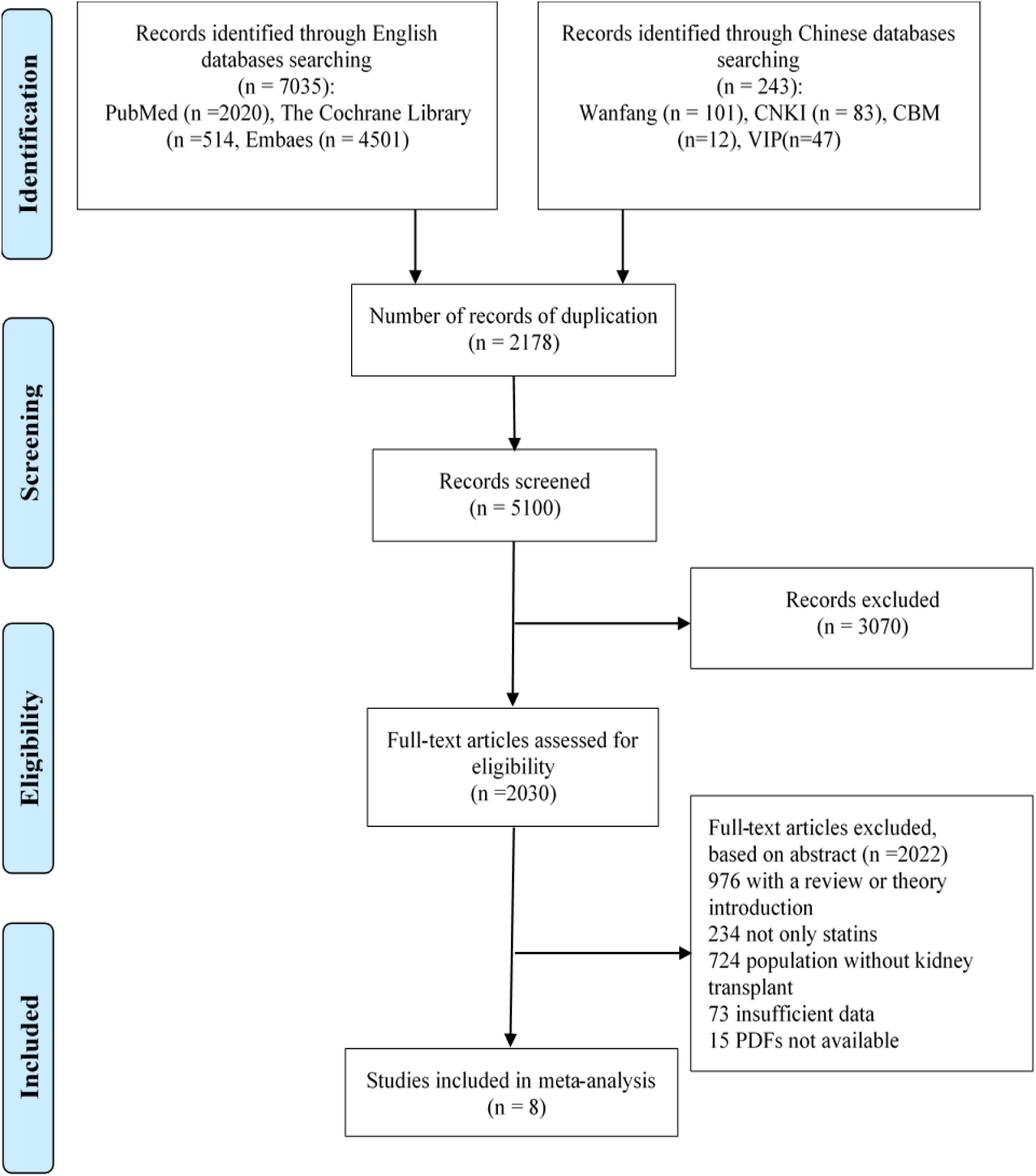
shows a flow chart of the selection process for eligible studies.

### 3.2. General characteristics of the studies included in the meta-analysis

All studies were published between 1995 and 2015. The number of participants ranged from 20 to 229. The treatment duration was as follows: one study had 4-week treatment 12, one study had 12-week treatment 16, three studies with 3-week treatment duration 10, 13, 17, and three studies had 6-week treatment duration 9, 14, 15. All participants underwent kidney transplantation. In all studies, patients received statins therapy. The statins prescribed at different clinical dosages ranging from 10 to 80 mg including, fluvastatin, simvastatin, pravastatin and atorvastatin. The follow-up duration was reported in all studies. The general characteristics of each included study are listed in Table 1.

**Table 1.** Characteristics of the studies included in this meta-analysis

| First author's name | Year | Country | Intervention | time (months) | Dose (mg/d) | Number | Follow-up (months) | TC achieved (mmol±L) | LDL-C achieved (mmol±L) | HDL-C achieved (mmol±L) | TG achieved (mmol±L) |
| --- | --- | --- | --- | --- | --- | --- | --- | --- | --- | --- | --- |
| Katznelson | 1995 | America | pravastatin | 4 | 20 | 24 | 24 | 5.2 | 2.79 | 1.45 | 1.34 |
| Renders | 2001 | Germany | atorvastatin | 3 | 10 | 10 | 10 | 4.97 | 2.75 | 1.32 | 1.79 |
| Hausberg | 2001 | Germany | fluvastatin | 6 | 40 | 18 | 18 | 6.19 | 3.57 | 1.86 | 1.84 |
| Santos | 2001 | Brazil | simvastatin | 6 | 10 | 34 | 33 | 5.42 | 3.04 | 1.44 | 2.05 |
| Kasiske | 2001 | America | simvastatin | 3 | 10 | 53 | 53 | 5.41 | 2.82 | 1.4 | 2.54 |
| Holdaas | 2001 | Norway | fluvastatin | 3 | 40 | 182 | 122 | 5.47 | 3.06 | 1.4 | 2.04 |
| Serond | 2008 | Spain | fluvastatin | 6 | 80 | 45 | 39 | 5.2 | 3.1 | 1.7 | 1.7 |
| Yazbek | 2015 | Brazil | pravastatin | 12 | 10 | 61 | 51 | 4.16 | 2.26 | 1.17 | 1.77 |

### 3.3. Risk of bias

The summary of risk of bias in each study is presented in Fig. 2.

**Fig. 2.**
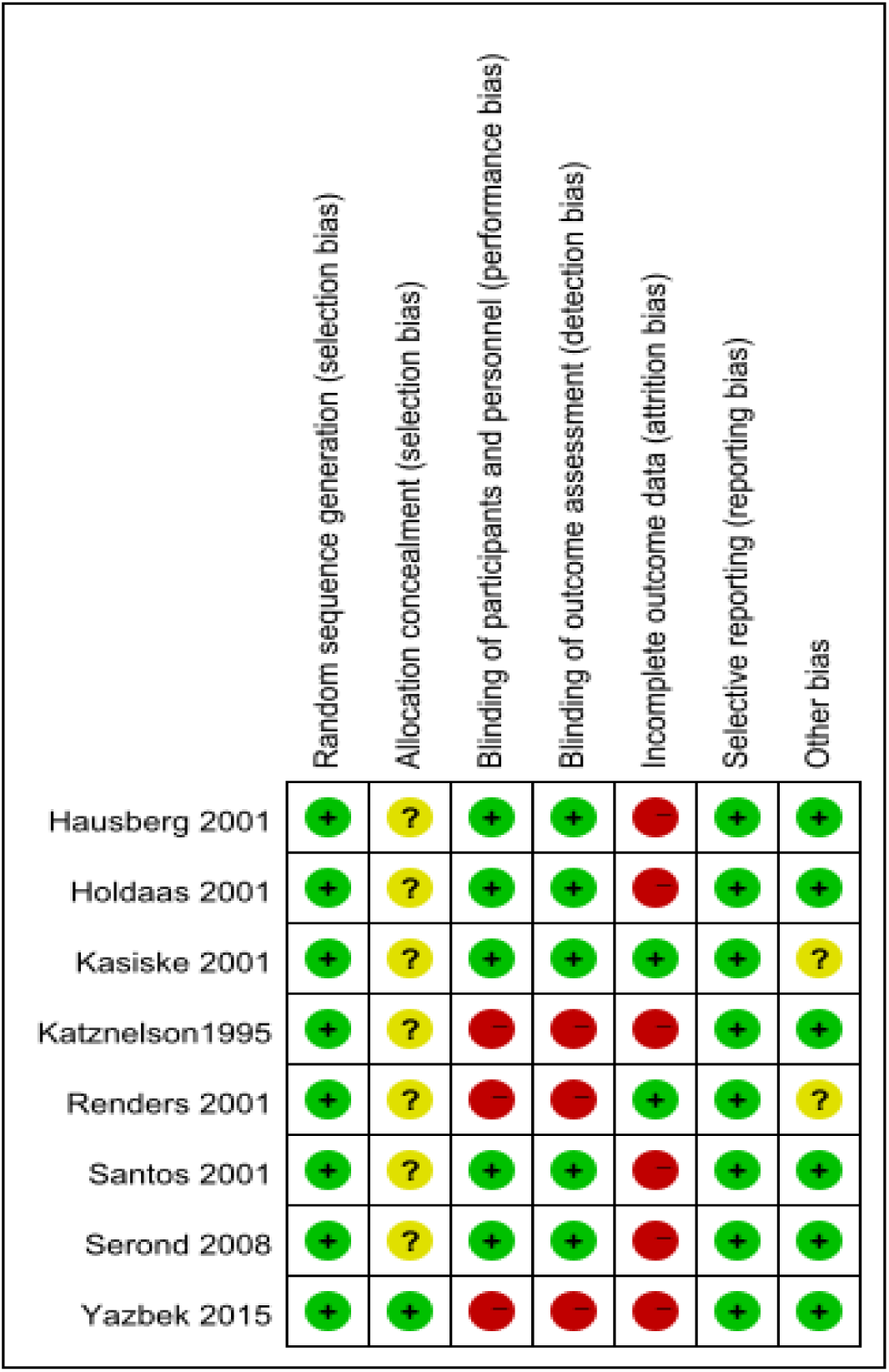
Summary of the risk of bias

### 3.4. Outcome analysis

The analysis identified high heterogeneity among the studies. Because the studies could not be combined, a random-effects model was used to pool the effect size for SMDs and 95% CIs for the analysis.

#### 3.4.1 The effects of statins on TC in KTRs

Eight trials reported the effect size of TC in KTRs (Table 1). The data show that the SMD was −2.90 with 95% CI:-4.52 to-1.27 compared to control group (p= 0.0005,Figure 3). The I^2^ value was 98%, suggesting the presence of heterogeneity. In addition, results revealed that statin therapy lowered TC levels.

**Fig. 3.**
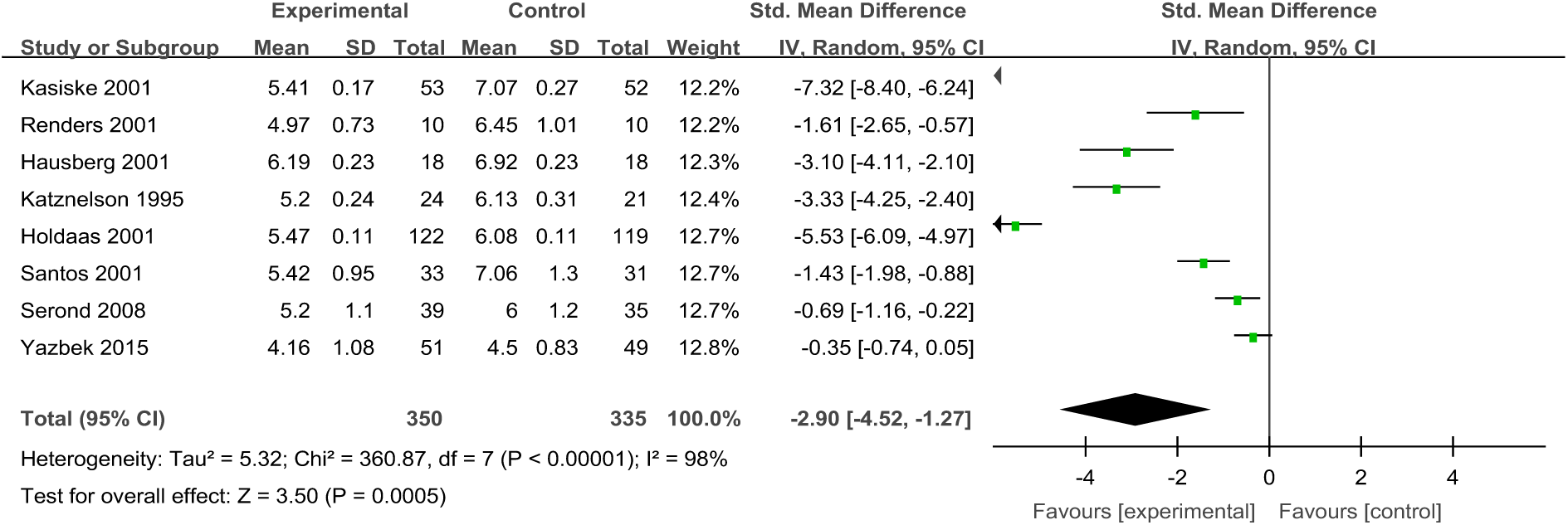
Forest plot of TC in KTRs

#### 3.4.2 The effect of statins on LDL-C in KTRs

Eight trials reported the effect size of LDL-C in KTRs who received statin therapy (Table 1). The analysis revealed that the SMD was −3.35 with 95% CI: −5.52 to −1.45 compared to control group (Figure 4). Although the level of heterogeneity was high (I^2^=98%; p<0.00001), the difference between the control group and the experimental group was statistically significant (p=0.0005). The results showed that statins therapy decreased the level of LDL-C compared to the controls.

**Fig. 4.**
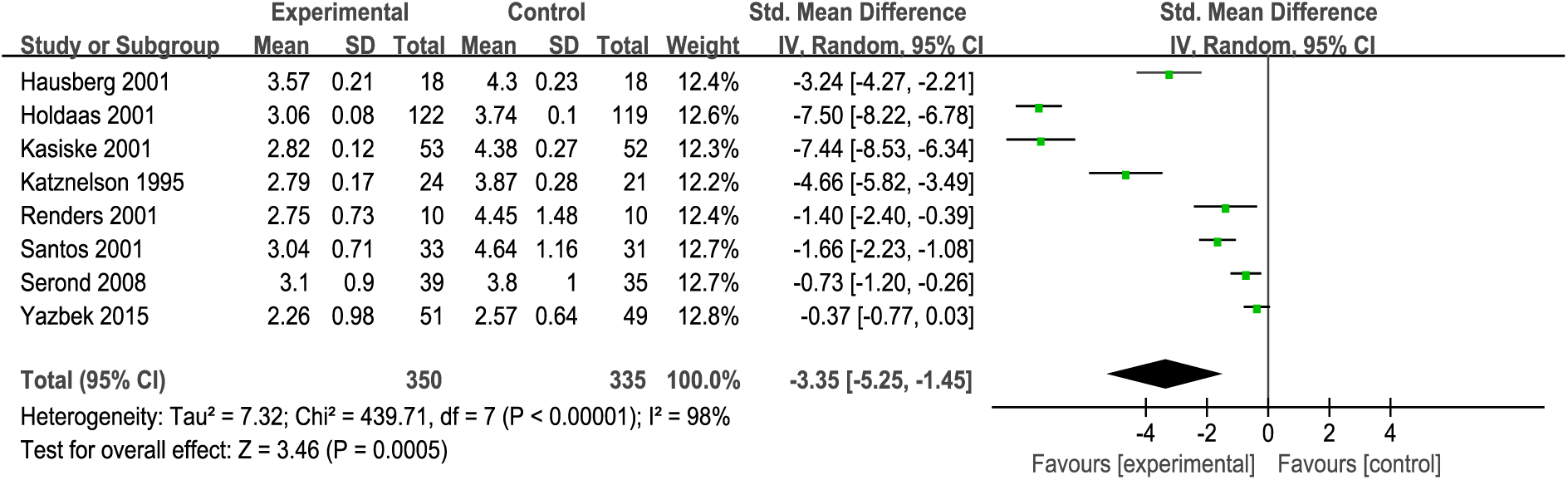
Forest plot of LDL-C in KTRs

#### 3.4.3 The effects of statins on HDL-C and TC in KTRs

Eight trials reported the level of HDL-C and TC (Table 1). The analysis revealed that there was no heterogeneity and none of them were no statistical significance. The SMD of HDL-C was −0.23 with 95% CI: −1.27 to 0.82 (p=0.67, Figure 5) and the SMD of TG was −0.39 with 95% CI: −1.45 to 0.66 (p=0.47, Figure 6). Results showed that statin therapy did not improve the profile of HDL-C or TG in KTRs.

**Fig. 6.**
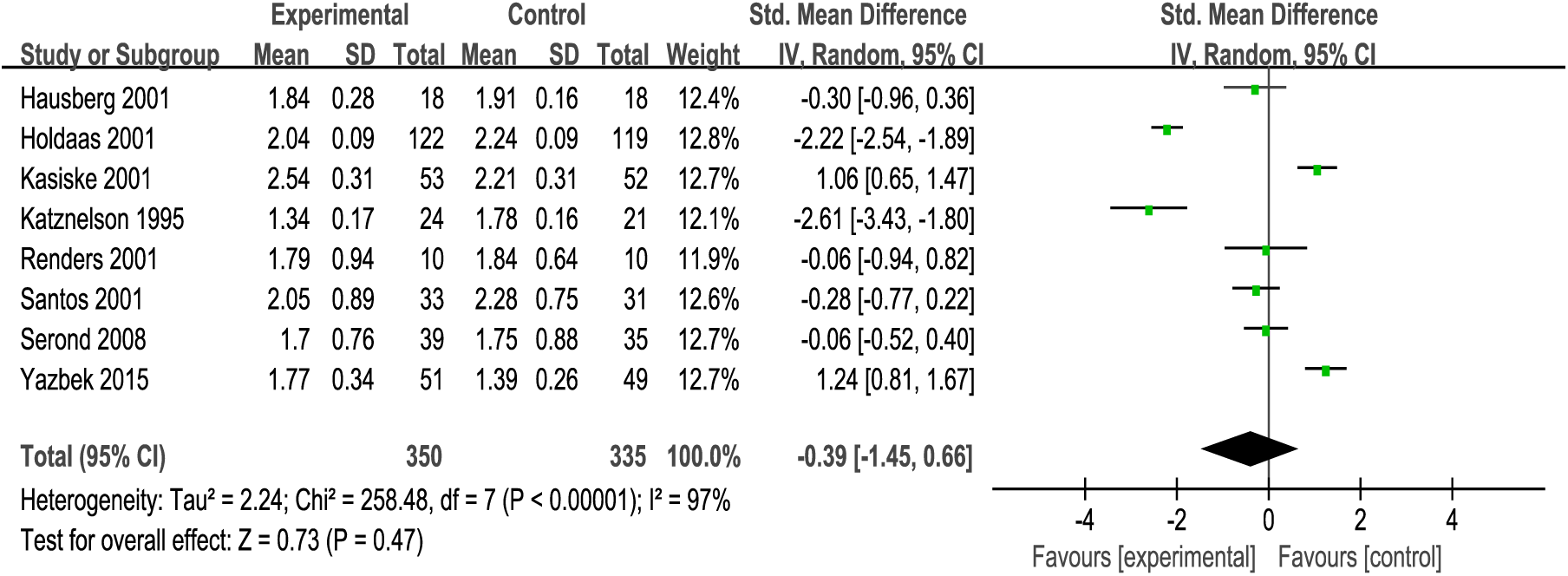
Forest plot of TG in KTRs

**Fig. 5.**
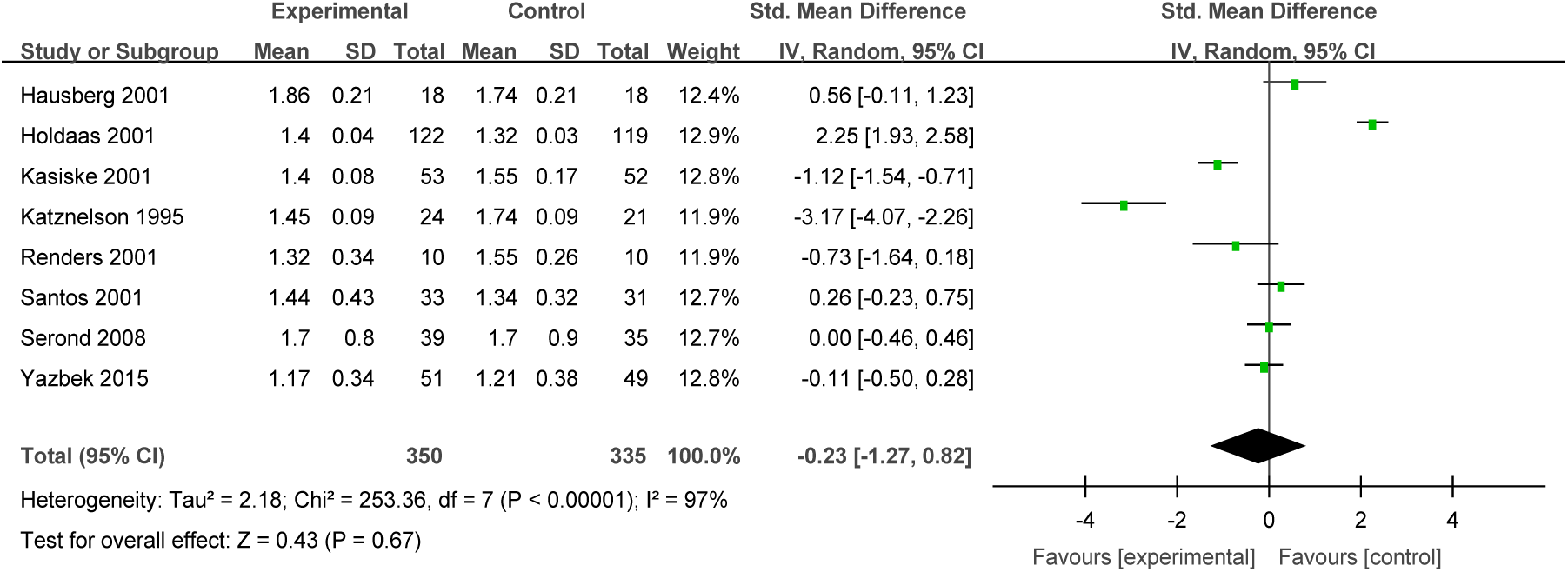
Forest plot of HDL-C in KTRs

### 3.5. Sensitivity analysis

Each study was individually excluded to determine the impact of each study on the outcomes. Because of the high heterogeneity, sensitivity analysis was performed for TC and LDL-C. Exclusion of studies by Holdaas et al. 10 and Kasiske et al. 11 decreased the heterogeneity level from 98% to 91% for TC, and from 98% to 93% for LDL-C. The conclusions made were not changed when studies with high risk of bias were omitted.

## 4. Discussion

### 4.1 Basic findings

This meta-analysis that performed on eight studies covering 350 kidney transplant patients who received statins and 335 kidney transplant patients as the control group. The aim of this study was to determine the clinical value of statins on lipid profile of KTRs. It was observed that statins therapy (including many types of statins) reduced serum levels of TC and LDL-C, which is consistent with previous reports5. However, statins had no significant effect on HDL-C and TG. Given the presence of risk of bias and the limited data, the findings of this analysis might not be conclusive. In addition, the effects of statins on cardiovascular events, cerebrovascular events, all-cause mortality and the specific characteristics could be analyzed. Large-scale and high-quality trials are required to evaluate the impact of statins on these parameters in KTRs.

So far, some large-scale studies have reported the effects of statins on CKD. The German Diabetes and Dialysis Study (4D)18 covering 1255 type 2 diabetes mellitus subjects who received maintenance hemodialysis, found that median level of LDL-C was reduced by 42% among patients receiving atorvastatin after four weeks of treatment. Another study evaluated the efficacy of rosuvastatin in 2776 patients undergoing regular hemodialysis (AURORA)19. They reported that administration of statins therapy for 3 months reduced the mean level of LDL-C by 43%. However, few studies have determined the influence of statins therapy on KTRs. In the ALERT trial 20, 2012 KTRs were enrolled and followed up for about five to six years. It was noted that the use of statins improved cholesterol levels in patients and increased the survival rate by 24% compared to patients who did not receive fluvastatin therapy. However, serum cholesterol level was not significantly associated with patient survival. Furthermore, some meta-analyses were performed to integrate the findings from these studies on KTRs.

To our knowledge, only a few meta-analyses have been designed to investigate the effects of statins on KTRs, but the conclusions from these studies are controversial. Palmer 5 argued that statins significantly reduced serum TC, LDL-C, HDL-C and TG, but they had no effect on overall mortality, stroke, kidney function, and toxicity outcomes in KTRs. Rostami 21 showed that the use of statins correlated independently with improved patient and graft survival after kidney transplantation. However, Messow 6 showed that it not convincing of lowering lipid about statins in patients after renal transplant. Krista 22argued that the use of statins cannot lower acute rejection risk after kidney transplantation. The evidence of use of statins in KTRs is insufficient. According to our meta-analysis, statins significantly lowered serum level of TC, and LDL-C, but had no effect on serum HDL-C and TG in patients who underwent kidney transplantation. It is likely that the nature and mechanisms of dyslipidemia and the features of CVD in KTRs lead to different conclusions. This implies that further studies should focus on the mechanisms of lipid disorders and the effect of statins therapy in KTRs.

In renal transplant patients, lipid disorders play a major role in renal dysfunction. This is associated with increased plasma levels of TC, LDL-C, VLDL-C, and TG as well as decreased plasma levels of HDL-C. Many factors can influence post-transplant lipid profile including age, genetic predisposition, obesity, reduced physical activity, diet and use of immunosuppressive agents (including calcineurin inhibitors, primarily cyclosporine, corticosteroids and mammalian target of rapamycin inhibitors)23. Interventions targeting dyslipidemia are advocated because it has the most fatal adverse effects among the risk factors of CVD. CVD is the most common cause of death in renal transplant patients24. Moreover, death with a functioning graft (with CVD the most common cause of death) is the overall most common cause of graft loss25. The proportion of the death with a functioning graft has increased over the 2 decades. And infections are the most leading causes that have not changed significantly26. In patients with end-stage renal disease, approximately 50% mortality is due to CVD and the incidence of CVD in KTRs is 4-6 times higher than in age-matched individuals 27, 28. Lipoprotein (a) is a modified form of LDL which regulates inflammation of the endothelium, thrombosis and binds macrophages to promote foam cell formation leading to the deposition of cholesterol in atherosclerotic plaques. In this way, it aggravates tissue injury and increases the risk of atherosclerosis and CVD-related morbidity and mortality in KTRs 29, 30. Therefore, future investigations are needed to develop interventions that manage lipid abnormalities.

Several agents have been designed to regulate lipid profile such as statins, niacin, fibrates, ezetimibe, fish oil/omega-3, bile acid resins, and PCSK9 inhibitors 31, 32 33, 34. So far, a number of studies have confirmed that statins are effective in lowering serum lipids and improving the cardiovascular risk compared to all other classes of medicines used in renal transplant patients 35.. Thus, both the kidney disease improving global outcomes (KDIGO) work group and European best practice guidelines for renal transplantation recommend the use of statins in adults who have received kidney transplant 36-38. In the past few years, several studies have identified the benefits of statins on survival of patients and reducing the risk of cardiovascular complications in KTRs. For instance, a study 39 found that KTRs treated with statins have a 24% higher survival rate than patients who do not receive statins. Another study 40 reported that that used of statins therapy in patients treated with tacrolimus after kidney transplant significantly lowered the risk of major adverse cardiovascular events. Recently, Michael 41 also found that a statistically significant survival benefit associated with the use of statins in a long-term follow-up retrospective study involving 687 ESRD patients who eventually underwent kidney transplant. Compelling evidence indicates that statins can be used to lower serum cholesterol (mainly aiming at LDL-C) after exercise and a low-fat diet has no effect on improving hypercholesterolemia. However, immunosuppressive agents are essential for KTRs. This is followed by adverse reactions, such as lipid metabolism disorders. Kidney transplantation and the use of immunosuppressive agents also contribute to lipid disorders42. For example, the use of chronic corticosteroid is associated with increases in TC, TG, and HDL-C43. Therefore, the effect of statins is more significant in KTRs. In addition to the cholesterol lowering effects, statins also exert anti-inflammatory, anti-proliferative effects, plaque-stabilizing capacities, improving endothelial function, antioxidant effects, immunosuppressive effects, regulate monocyte recruitment, matrix deposition and renal hemodynamics 5, 44-46. In addition, the anti-inflammatory effect of statins should not be ignored. Results from the studies demonstrate the ability of statins that improve graft outcome in the first year post-transplantation and inhibit transplant rejection47 48. Besides statins established role in the management of lipid profile, it may be closely attributed to the various anti-inflammatory effect of statins. It was proved that statins can impinge on chemokine and inflammatory cytokines release, on prostaglandin expression and on effector phagocytic function of monocytes49, 50. Moreover, statins also could ameliorate complement-mediated vascular damage that is vital to the initiation and perpetuation of inflammation51. Therefore, not only lipid-lowering but also anti-inflammatory is beneficial for reduction of rejection episodes that are associated with enhancing graft longevity. As the rate-limiting step in cholesterol biosynthesis, HMG-CoA reductase is a major inhibition target through which statins reduce intracellular cholesterol in the liver and stimulate the expression of LDL receptors, increase receptor-mediated endocytosis of LDL; thereby lowering serum LDL and TC. It should be noted that there are other effects that mildly reduce TG and modestly elevate HDL. We’d better take into consideration that dyslipidemia may persist along time after renal transplantation, though statin treatment is persistent 52.So it’s necessary to evaluate cholesterol levels regularly (including TC, LDL-C, HDL-C and TG). As we all know, the methods for LDL-C measurement is progressing such as ultracentrifugation, the Friedewald equation and the Martin–Hopkins equation53, 54. Although these methods of LDL-C are alternative, the results are not very different. Moreover, we did choose the SMDs to eliminate the differences in some aspects.

Before prescribing statins to decrease lipid profile, secondary factors that elevate lipids such as nephrotic syndrome, hypothyroidism, diabetes mellitus, excessive alcohol intake, chronic liver disease and other medication-induced dyslipidemias should be excluded 55.

### 4.2 Strengths and limitations of the study

This meta-analysis has some potential limitations. Firstly, some factors that influence heterogeneity such as race, gender, baseline serum lipid levels, use of different statins, different dosage and durations of statins therapy, duration of follow-up and the nature of patient population were not analyzed. Secondly, only pooling eight studies were used to analyze the effects of statins on KTRs subjects. Thirdly, it is regretful that we can’t get the eligible studies from the Chinese database.

## 5. Conclusion

Statins therapy significantly improves lipid profile (such as decreasing TG and LDL-C) in patients with KTRs. Therefore, statins should be prescribed to KTRs. However, more high-quality, large scale randomized controlled trials are needed to explore the effect of statins therapy in patients with dyslipidemia after renal transplantation. In addition, the underlying mechanisms responsible for dyslipidemia such as the type of immunosuppressant used requires further investigation. Furthermore, the ongoing clinical trials should evaluate the adverse events of statins.

## Data Availability

Data types can be obtained from the corresponding author on request to support the results of this study.

## Ethical approval

None.

## Acknowledgment

We would like to thank Qiang Zhang for his valuable suggestions and statistical review.

